# AI-Driven Early Detection of Polycystic Ovary Syndrome via Follicle Count

**DOI:** 10.64898/2026.09.01.26361974

**Authors:** Divya Thota, Aishwarya Mahesha, Mohammed Fazil Khasim, Khyathi Priya Kethineni, Bhavana Pothireddygari, Bahareh Rahmani

## Abstract

Polycystic Ovary Syndrome is a common endocrine disorder characterized by ovulatory dysfunction, hyperandrogenism, and/or polycystic ovarian morphology, with significant reproductive and metabolic consequences. Due to heterogeneous symptom profiles, Polycystic Ovary Syndrome is frequently underdiagnosed or diagnosed late. In this study, we develop machine learning models for early Polycystic Ovary Syndrome prediction using a structured clinical dataset with 42 features and 542 patient records. After data cleaning and normalization, correlation-based feature selection was applied to retain the most predictive variables. Multiple models were trained and evaluated, including Logistic Regression, Decision Tree, KNN, and Random Forest. Results demonstrate that Random Forest achieves the best overall performance (approximately 88% accuracy), suggesting that ensemble models can effectively capture non-linear feature interactions in clinical data. We also contextualize findings with international clinical guidance and recent work on explainable and clinically applicable Polycystic Ovary Syndrome prediction systems.

## 1. Introduction

Polycystic Ovary Syndrome is a prevalent and heterogeneous endocrine disorder affecting women of reproductive age [1,2]. International guidance emphasizes standardized diagnostic pathways and long-term risk assessment due to associated metabolic and cardiovascular complications [3,4]. Polycystic Ovary Syndrome prevalence estimates vary with diagnostic criteria; systematic reviews suggest prevalence around 6% under NIH criteria and closer to 10% under Rotterdam criteria subsets [5,6].

In clinical practice, Polycystic Ovary Syndrome is important not only because of reproductive symptoms (irregular cycles, infertility, hyperandrogenic signs) but also because of long-term comorbidities such as insulin resistance, dyslipidemia, hypertension, and increased cardiometabolic risk [7–9]. Multiple guidelines (e.g., Endocrine Society and AACE/ACE/AE-Polycystic Ovary Syndrome statements) highlight the need for integrated screening and management of both reproductive and metabolic domains [10,11].

Diagnosis remains challenging because presentation differs across phenotypes and life stages, and diagnostic criteria have evolved over time [2,12,13]. Widely adopted standards, such as the Rotterdam criteria, define Polycystic Ovary Syndrome by the presence of at least two of three features—ovulatory dysfunction, hyperandrogenism, and polycystic ovarian morphology—after excluding related disorders [12]. The International Evidence-Based Polycystic Ovary Syndrome Guidelines (2018, 2023) provide structured recommendations for assessment and management and note that ultrasound is not always required when irregular cycles and hyperandrogenism are present [3,4].

Machine learning approaches can integrate multidimensional clinical variables to identify complex patterns and provide screening support. Systematic review evidence suggests that AI/ML methods can achieve strong diagnostic performance, while emphasizing the importance of standardized diagnostic criteria, rigorous evaluation design, and external validation [14]. This paper presents a machine-learning-based Polycystic Ovary Syndrome prediction workflow and compares common models under consistent preprocessing and evaluation.

### Clinical criteria and epidemiology

Polycystic Ovary Syndrome diagnostic criteria and phenotypic variation have been extensively discussed in major reviews and consensus statements [2,6,12,13,15]. Prevalence estimates vary by region and diagnostic framework, as demonstrated in meta-analyses [5]. Guidelines also highlight patient experiences, including delayed diagnosis and dissatisfaction due to lack of information [16].

### Role of ultrasound and biomarkers

Ultrasound evidence of polycystic ovarian morphology remains a key diagnostic component, though optimal markers and thresholds vary by imaging modality and technical constraints. Recent evidence supports follicle number per ovary (FNPO) as a strong diagnostic marker, while ovarian volume and follicle number per section serve as alternatives when full follicle counts are not feasible [3,17]. Anti-Müllerian hormone (AMH) has also been actively studied as a complementary biomarker; meta-analyses informing the 2023 guideline highlight its utility and limitations such as assay variability, age, phenotype, and BMI dependence—supporting its use as an adjunct rather than a standalone diagnostic criterion [18].

### AI/ML for POS prediction

Recent work includes feature-selection-driven tabular ML pipelines [19,20], explainable ensemble approaches [21], and stacked or balanced frameworks incorporating advanced feature selection [22]. Imaging-based approaches using ultrasound data with CNNs and ensemble models have also demonstrated high diagnostic accuracy [23,24]. A systematic review in *Frontiers in Endocrinology* summarizes AI/ML applications for Polycystic Ovary Syndrome diagnosis and classification, emphasizing standardized diagnostic definitions and transparent reporting [14]. More recent multicenter, interpretable frameworks using gradient-boosted models and SHAP analysis demonstrate strong discrimination and transparent assessment of feature contributions [25]. Explainability methods such as SHAP provide a unified framework for interpreting complex predictive models [26].

## 2. Data Description

The dataset used in this project is a structured Polycystic Ovary Syndrome dataset with 542 patient records and dozens of clinical/physical parameters (e.g., BMI, hormone levels, follicle counts) commonly distributed for research and benchmarking [28]. The target label indicates Polycystic Ovary Syndrome status (Y/N).

The features include a mixture of continuous variables (e.g., hormone concentrations, anthropometrics) and binary/categorical indicators (e.g., symptoms such as acne/hirsutism indicators, cycle regularity, lifestyle-related features). This mixture motivates preprocessing steps such as scaling/normalization and careful feature selection to ensure models are not dominated by variables with larger numeric ranges.

## 2. Methodology

### Data Reading and Cleaning

The dataset was cleaned by handling missing values and normalizing feature scales to reduce dominance by large-range numeric features. Where missingness occurred, records and features were reviewed to ensure that basic model training requirements (consistent input dimensions and valid numeric representations) were satisfied.

### Train/Test Split and Reproducibility

A train-test split was used to evaluate generalization. The training set was used for model fitting and the test set was reserved for evaluation. Reproducibility is supported by using deterministic random seeds during splitting and model training when available.

### Feature Selection

Correlation analysis was applied to identify the most relevant predictors. The top correlated predictors typically include follicle counts (left/right), cycle regularity, signs of hyperandrogenism, and related clinical indicators, consistent with diagnostic criteria and guideline emphasis on ovarian morphology and hyperandrogenism.

### Exploratory Visualizations

Histograms, density plots, and box plots were used to understand distributions and outliers. These visualizations help identify skewness, heavy tails, and anomalous values that may influence model stability, particularly in distance-based methods like KNN.

### Model Training and Evaluation

We trained and evaluated Logistic Regression, Decision Tree, KNN, and Random Forest [5]. Performance was assessed using accuracy and confusion matrix-based measures; for clinical decision-support settings, additional metrics such as precision, recall, and F1-score. Figure 1 shows the top correlated features with Polycystic Ovary Syndrome labels based on correlation analysis.

**Figure 1.**
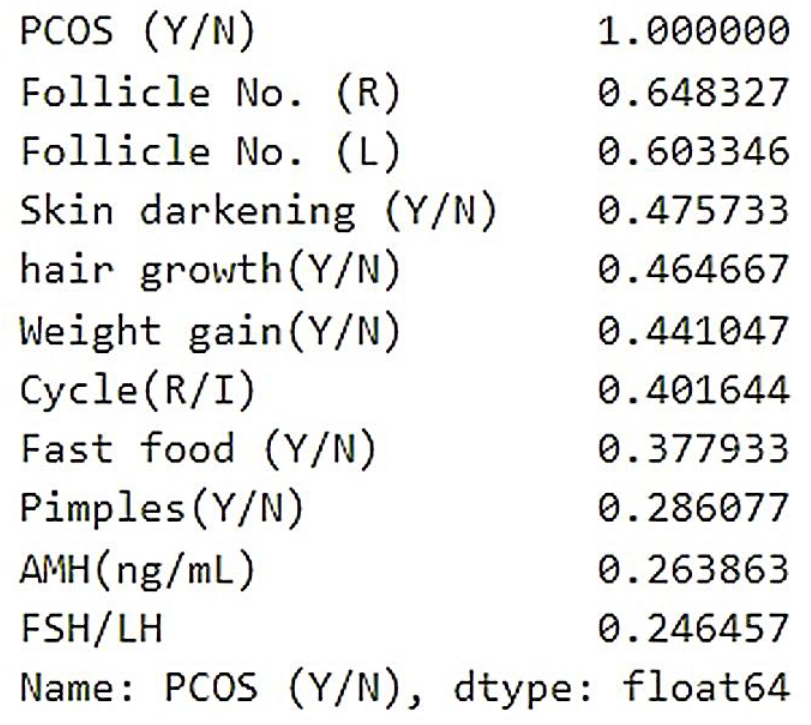
Top correlated features with POLYCYSTIC OVARY SYNDROME labels

## 4. Results

### Logistic Regression

Logistic Regression produced a strong baseline accuracy (about 80%), consistent with prior tabular Polycystic Ovary Syndrome prediction studies. This model provides a useful reference point due to its relative interpretability and stability.

### Decision Tree

A Decision Tree achieved moderate accuracy and provides interpretability through rule thresholds. Although single trees can overfit, they offer human-readable decision logic that can be compared against clinical intuition. Figure 2 shows a decision tree with %79 accuracy.

**Figure 2.**
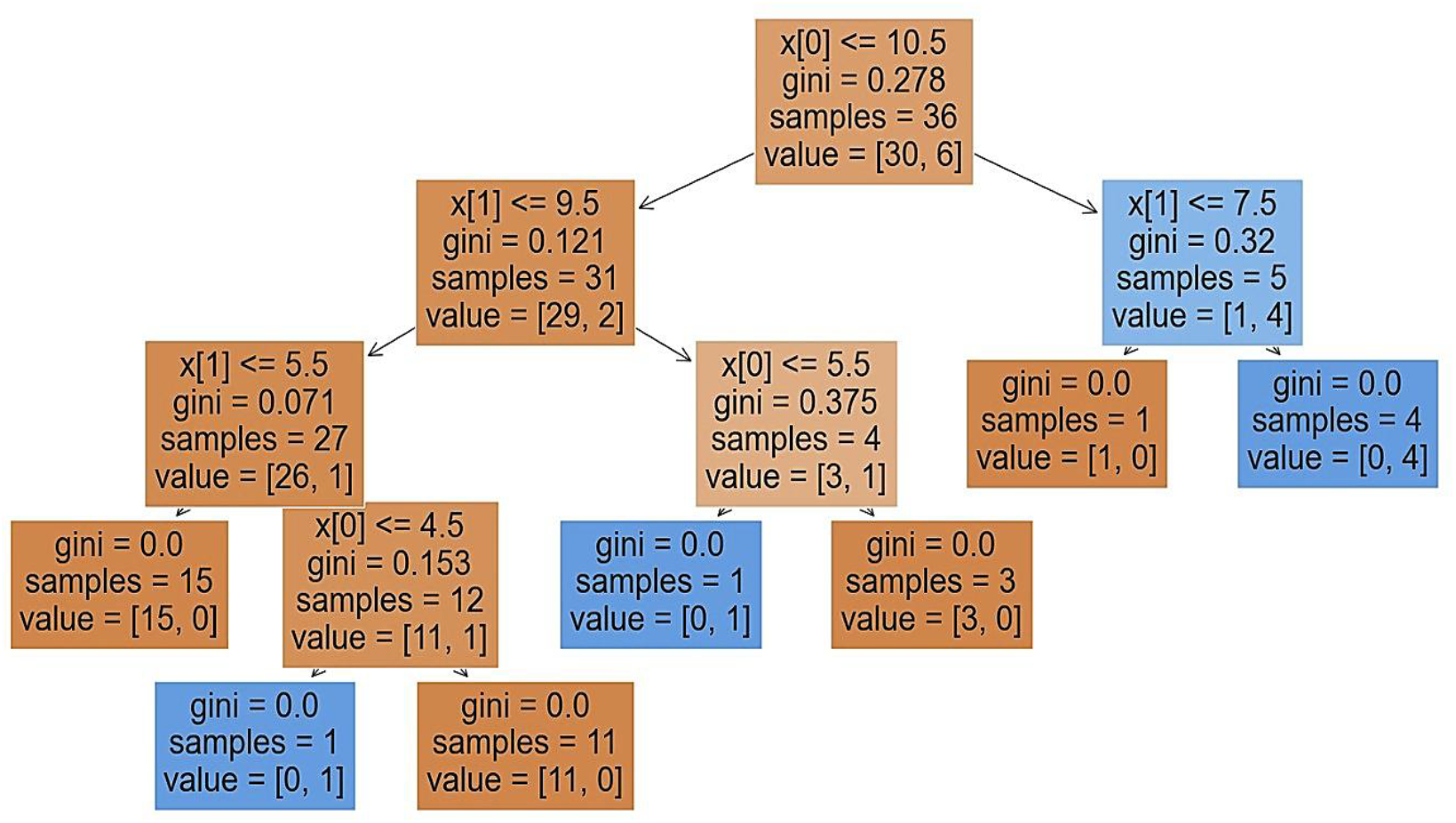
Decision tree visualization

### KNN (k = 4)

The elbow method suggested an effective choice; KNN achieved competitive accuracy. KNN performance depends on scaling and distance behavior, which motivates normalization and outlier review in preprocessing. Figure 3 shows the KNN graph with %83 accuracy.

**Figure 3.**
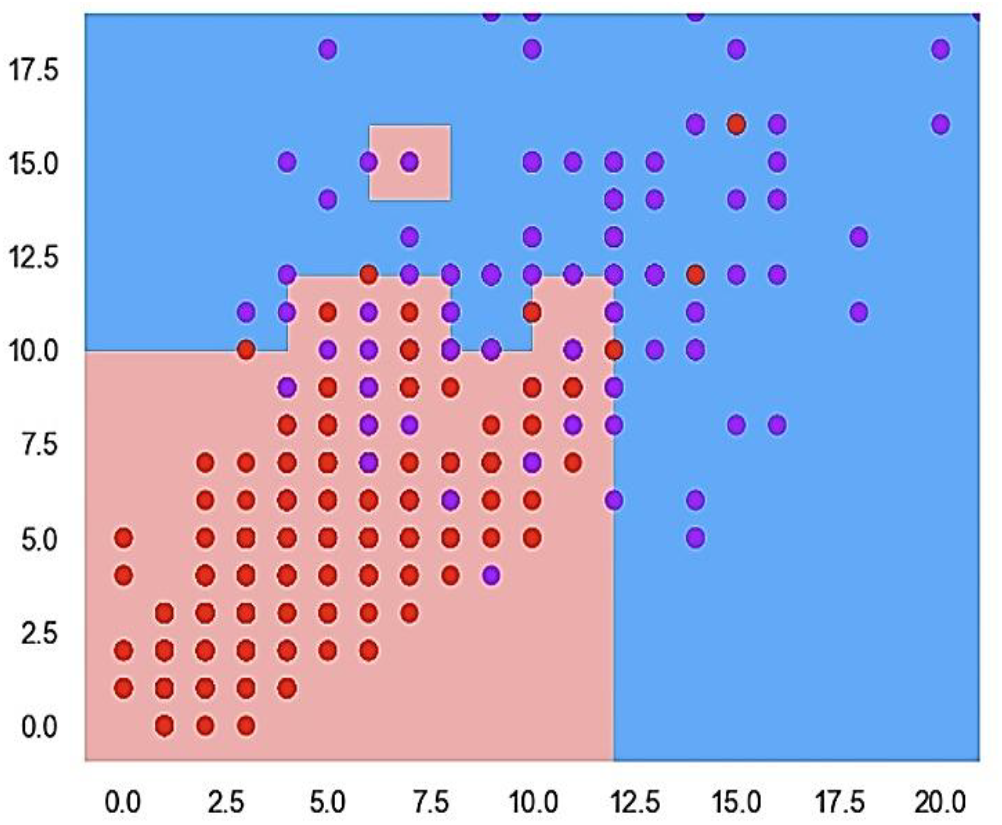
KNN visualization and performance

### Random Forest

Random Forest achieved the best overall performance (approximately 88% accuracy), consistent with literature where three ensembles often perform strongly on mixed clinical data. Random Forest can model non-linear interactions and reduce variance through bagging. Figure 4 shows the Random Forest confusion matrix, accuracy, and classification report. Three trees of Random Forest are shown in figure 5.

**Figure 4.**
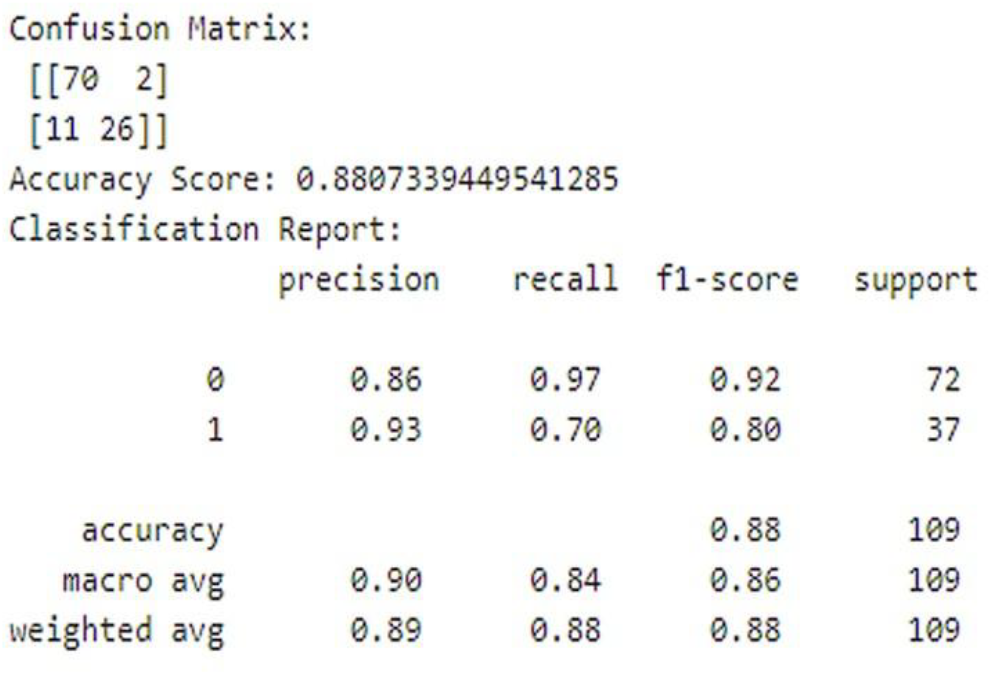
Random Forest confusion matrix, accuracy, and classification report

**Figure 5.**
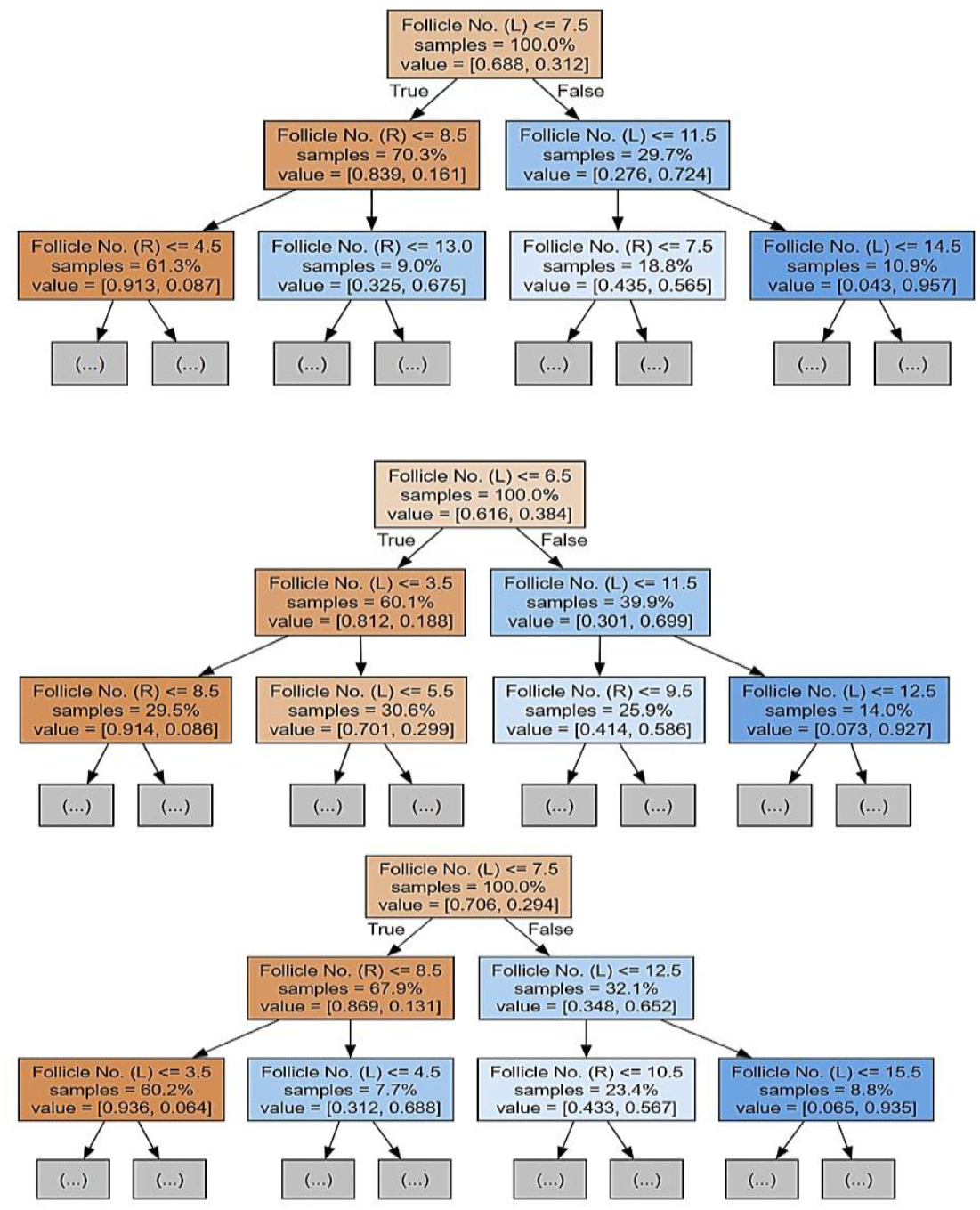
Sample trees of Random Forest

### Model Accuracy Comparison

Figure 6 summarizes test accuracy for each evaluated classifier. While accuracy is informative, future screening-oriented work should also emphasize sensitivity/recall for minimizing missed patients.

Based on Figure 7, Random Forest and KNN show the highest accuracy.

**Figure 7.**
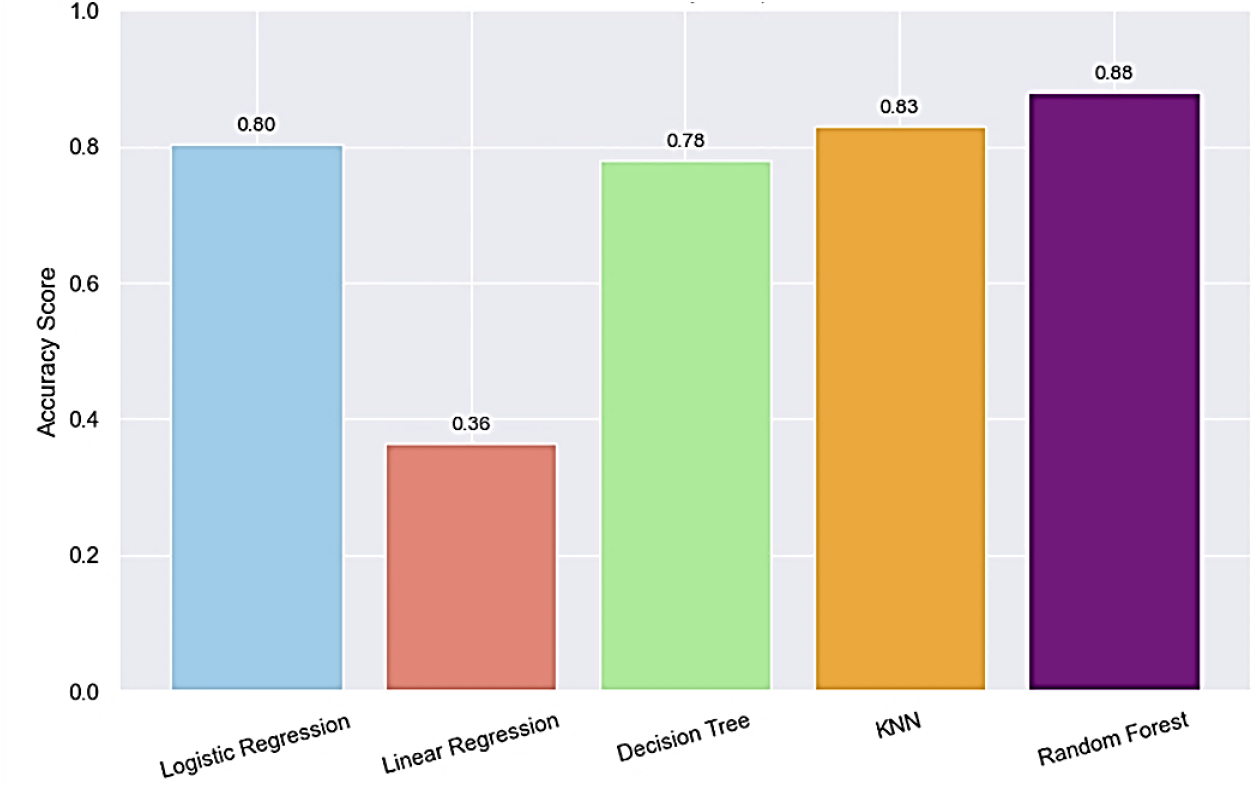
Model Comparison

## 5. Discussion

In this study, we evaluated multiple machine learning models for early prediction of Polycystic Ovary Syndrome using a structured clinical dataset comprising hormonal, anthropometric, and symptom-based features. Across all evaluated models, ensemble-based and instance-based approaches outperformed simpler linear classifiers, with Random Forest achieving the highest overall accuracy (~88%). These findings are consistent with prior work demonstrating the strength of tree-based ensembles in handling heterogeneous clinical data with non-linear interactions and mixed feature types [5,19–23].

The superior performance of Random Forest likely reflects its ability to capture complex interactions among clinical indicators that mirror real-world diagnostic reasoning. Features such as follicle counts, menstrual irregularity, and signs of hyperandrogenism were among the most strongly correlated predictors, aligning well with established diagnostic frameworks such as the Rotterdam criteria and international Polycystic Ovary Syndrome guidelines [2–4,12]. This concordance between data-driven feature importance and clinical criteria supports the face validity of the proposed modeling approach.

Logistic Regression provided a strong and interpretable baseline (~80% accuracy), highlighting that a substantial portion of Polycystic Ovary Syndrome risk can be explained by additive effects of key clinical variables. However, its comparatively lower performance suggests that purely linear decision boundaries may be insufficient to fully capture the heterogeneous phenotypes of Polycystic Ovary Syndrome described in epidemiological and mechanistic studies [6,11,14]. Decision Trees offered intuitive rule-based interpretations but showed reduced generalization, likely due to sensitivity to noise and overfitting in limited sample sizes.

KNN demonstrated competitive performance when appropriate normalization was applied, reinforcing the importance of careful preprocessing in distance-based models. However, KNN’s reliance on local neighborhood structure and sensitivity to outliers may limit its robustness in larger or more diverse clinical cohorts, particularly when missingness or measurement variability is present.

From a clinical perspective, these results support the role of machine learning as a screening and decision-support tool, rather than a replacement for formal diagnostic criteria. International guidelines emphasize that Polycystic Ovary Syndrome diagnosis requires exclusion of alternative etiologies and consideration of patient context, which cannot be fully captured in a single dataset [3,4]. Nevertheless, ML-based tools may help identify high-risk individuals earlier, potentially reducing diagnostic delays that are commonly reported by patients [16].

Importantly, while overall accuracy was used as the primary comparison metric in this study, future clinically oriented implementations should prioritize sensitivity and recall, particularly to minimize missed Polycystic Ovary Syndrome cases. This is especially relevant in screening settings where false negatives may delay appropriate follow-up and management. Explainability methods such as SHAP, which have been successfully applied in recent multicenter studies [25–27], represent a natural next step to enhance transparency and clinician trust.

Several limitations should be acknowledged. First, the dataset size, while comparable to many benchmarks Polycystic Ovary Syndrome studies, limits external generalizability. Second, the dataset represents a structured research cohort rather than real-world electronic health record (EHR) data, which may include greater noise, missingness, and population heterogeneity. Third, imaging-based and longitudinal data were not incorporated, despite growing evidence supporting the integration of ultrasound markers and temporal clinical trajectories [17,20,23,24].

## 6. Conclusion

This study demonstrates that machine learning models, particularly ensemble methods such as Random Forest, can effectively predict Polycystic Ovary Syndrome using routinely collected clinical features. By achieving approximately 88% accuracy, the proposed approach highlights the potential of data-driven models to support earlier identification of Polycystic Ovary Syndrome in settings where diagnostic uncertainty and delayed recognition remain common.

Our findings are consistent with international diagnostic guidance and prior AI/ML literature, reinforcing that key reproductive and hyperandrogenic features remain central to Polycystic Ovary Syndrome prediction, even in automated frameworks. Importantly, the comparative evaluation of multiple models under consistent preprocessing provides practical insight into model selection for clinical decision-support applications.

Future work will focus on external validation in independent and multi-institutional cohorts, incorporation of explainable AI techniques to enhance clinical interpretability, and expansion to multimodal data sources such as ultrasound imaging and longitudinal follow-up. Ultimately, integrating machine learning–based screening tools into clinical workflows may help reduce diagnostic delays, support personalized risk stratification, and improve long-term reproductive and metabolic outcomes for individuals with Polycystic Ovary Syndrome.

## Data Availability

The dataset used in this project is a structured Polycystic Ovary Syndrome dataset with 542 patient records and dozens of clinical/physical parameters (e.g., BMI, hormone levels, follicle counts) commonly distributed for research and benchmarking. The target label indicates Polycystic Ovary Syndrome status (Y/N).
The features include a mixture of continuous variables (e.g., hormone concentrations, anthropometrics) and binary/categorical indicators (e.g., symptoms such as acne/hirsutism indicators, cycle regularity, lifestyle-related features). This mixture motivates preprocessing steps such as scaling/normalization and careful feature selection to ensure models are not dominated by variables with larger numeric ranges

https://www.kaggle.com/datasets/prasoonkottarathil/polycystic-ovary-syndrome-pcos

## Data Availability

The data are available in: https://www.kaggle.com/datasets/prasoonkottarathil/polycystic-ovary-syndrome-Polycystic Ovary Syndrome

## Author Contribution

Divya Thota: Write the original paper, Methodology, Analysis.

Aishwarya Mahesha: Visualization, Methodology, Analysis.

Mohammed Fazil Khasim: Validation, Programming, Analysis.

Khyathi Priya Kethineni: Data collection, Literature Review, Visualization.

Bhavana Pothi Reddy Gari: Data curation, Visualization.

Bahareh Rahmani, Project’s supervision, Admin, Review the paper, Validation.

## References

[1] World Health Organization, “Polycystic ovary syndrome (POLYCYSTIC OVARY SYNDROME),” https://www.who.int/news-room/fact-sheets/detail/polycystic-ovary-syndrome, 2025, accessed: 2026-01-14.

[2] The Rotterdam ESHRE/ASRM-Sponsored POLYCYSTIC OVARY SYNDROME Consensus Workshop Group, “Revised 2003 consensus on diagnostic criteria and long-term health risks related to polycystic ovary syndrome (POLYCYSTIC OVARY SYNDROME),” Human Reproduction, vol. 19, no. 1, pp. 41–47, 2004.

[3] H. J. Teede, C. T. Tay, J. J. E. Laven, A. Dokras, L. J. Moran, T. T. Piltonen, M. F. Costello, J. Boivin et al., “Recommendations from the 2023 international evidence-based guideline for the assessment and management of polycystic ovary syndrome,” The Journal of Clinical Endocrinology & Metabolism, vol. 108, no. 10, pp. 2447–2469, 2023.

[4] M. Gibson-Helm, H. Teede, A. Dunaif, and A. Dokras, “Delayed diagnosis and a lack of information associated with dissatisfaction in women with polycystic ovary syndrome,” The Journal of Clinical Endocrinology & Metabolism, vol. 102, no. 2, pp. 604–612, 2017.

[5] L. Breiman, “Random forests,” Machine Learning, vol. 45, no. 1, pp. 5–32, 2001.

[6] R. S. Legro, S. A. Arslanian, D. A. Ehrmann, K. M. Hoeger, M. H. Murad, R. Pasquali, and C. K. Welt, “Diagnosis and treatment of polycystic ovary syndrome: An Endocrine Society clinical practice guideline,” The Journal of Clinical Endocrinology & Metabolism, vol. 98, no. 12, pp. 4565–4592, 2013.

[7] N. F. Goodman, R. H. Cobin, W. Futterweit, C. J. Glueck, R. S. Legro, and E. Carmina, “American Association of Clinical Endocrinologists, American College of Endocrinology, and Androgen Excess and POLYCYSTIC OVARY SYNDROME Society disease state clinical review: Guide to the best practices in the evaluation and treatment of polycystic ovary syndrome,” Endocrine Practice, 2015.

[8] H. Elmannai, N. El-Rashidy, I. Mashal et al., “Polycystic ovary syndrome detection machine learning model based on optimized feature selection and explainable artificial intelligence,” Diagnostics, vol. 13, no. 8, p. 1506, 2023.

[9] U. Mahesswari and U. Maheswari, “SmartScanPOLYCYSTIC OVARY SYNDROME: A feature-driven approach to cutting-edge prediction of polycystic ovary syndrome using machine learning and explainable artificial intelligence,” Heliyon, vol. 10, no. 20, p. e39205, 2024.

[10] F. J. Barrera et al., “Application of machine learning and artificial intelligence in the diagnosis and classification of polycystic ovarian syndrome: A systematic review,” Frontiers in Endocrinology, 2023.

[11] D. A. Dumesic, S. E. Oberfield, E. Stener-Victorin, J. C. Marshall, J. S. Laven, and R. S. Legro, “Scientific statement on the diagnostic criteria, epidemiology, pathophysiology, and molecular genetics of polycystic ovary syndrome,” Endocrine Reviews, vol. 36, no. 5, pp. 487–525, 2015.

[12] H. J. Teede, M. L. Misso, M. F. Costello, A. Dokras, J. Laven, L. Moran, T. Piltonen, R. J. Norman, and I. P. Network, “Recommendations from the international evidence-based guideline for the assessment and management of polycystic ovary syndrome,” Human Reproduction, vol. 33, no. 9, pp. 1602–1618, 2018.

[13] G. Bozdag, S. Mumusoglu, D. Zengin, E. Karabulut, and B. O. Yildiz, “The prevalence and phenotypic features of polycystic ovary syndrome: A systematic review and meta-analysis,” Human Reproduction, vol. 31, no. 12, pp. 2841–2855, 2016.

[14] D. Lizneva, L. Suturina, W. Walker, S. Brakta, L. Gavrilova-Jordan, and R. Azziz, “Criteria, prevalence, and phenotypes of polycystic ovary syndrome,” Fertility and Sterility, vol. 106, no. 1, pp. 6–15, 2016.

[15] C. T. Tay et al., “2023 international evidence-based polycystic ovary syndrome guideline: Cardiometabolic considerations,” Journal of the American Heart Association, 2024.

[16] P. Dubey et al., “Polycystic ovary syndrome, insulin resistance, and cardiovascular disease,” Current Opinion in Endocrinology, Diabetes and Obesity, 2024, PubMed: 38568339. [Online]. Available: https://pubmed.ncbi.nlm.nih.gov/38568339/

[17] K. B. Studen and M. Pfeifer, “Cardiometabolic risk in polycystic ovary syndrome,” Endocrine Connections, vol. 7, no. 7, pp. R238–R251, 2018.

[18] R. Azziz, E. Carmina, D. Dewailly et al., “Criteria for defining polycystic ovary syndrome as a predominantly hyperandrogenic syndrome: An Androgen Excess Society guideline,” The Journal of Clinical Endocrinology & Metabolism, vol. 91, no. 11, pp. 4237–4245, 2006.

[19] R. Azziz, E. Carmina, D. Dewailly et al., “The Androgen Excess and POLYCYSTIC OVARY SYNDROME Society criteria for the polycystic ovary syndrome: The complete task force report,” Fertility and Sterility, vol. 91, no. 2, pp. 456–488, 2009.

[20] J. Peña et al., “Ultrasonographic criteria in the diagnosis of polycystic ovary syndrome: A diagnostic meta-analysis,” Human Reproduction Update, 2024, PubMed: 37804097. [Online]. Available: https://pubmed.ncbi.nlm.nih.gov/37804097/

[21] K. van der Ham et al., “Anti-Müllerian hormone as a diagnostic biomarker for polycystic ovary syndrome: A meta-analysis,” Fertility and Sterility, 2024, PubMed: 38944177. [Online]. Available: https://pubmed.ncbi.nlm.nih.gov/38944177/

[22] B. Panjwani, J. Yadav, V. Mohan, N. Agarwal, and S. Agarwal, “Optimized machine learning for the early detection of polycystic ovary syndrome in women,” Sensors, vol. 25, no. 4, p. 1166, 2025.

[23] H. M. Emara, W. El-Shafai, N. F. Soliman, A. D. Algarni, R. Alkanhel, and F. E. Abd El-Samie, “A stacked learning framework for accurate classification of polycystic ovary syndrome with advanced data balancing and feature selection techniques,” Frontiers in Physiology, vol. 16, p. 1435036, 2025.

[24] S. A. Suha and M. N. Islam, “An extended machine learning technique for polycystic ovary syndrome detection using ovary ultrasound image,” Scientific Reports, vol. 12, p. 17123, 2022.

[25] J. Kermanshahchi et al., “Development of a machine learning-based model for accurate identification of POLYCYSTIC OVARY SYNDROME pelvic ultrasound images,” 2024. [Online]. Available: https://pmc.ncbi.nlm.nih.gov/articles/PMC11338641/

[26] B. Yao et al., “Development and validation of an explainable machine learning and nomogram model for early detection and risk stratification of polycystic ovary syndrome: A multicenter study,” Frontiers in Endocrinology, 2025.

[27] S. M. Lundberg and S.-I. Lee, “A unified approach to interpreting model predictions,” in Advances in Neural Information Processing Systems (NeurIPS), 2017. [Online]. Available: https://dl.acm.org/doi/10.5555/3295222.3295230

[28] K. D. Contributor(s), “Polycystic ovary syndrome (POLYCYSTIC OVARY SYNDROME) dataset,” https://www.kaggle.com/datasets/prasoonkottarathil/polycystic-ovary-syndrome-PolycysticOvarySyndrome, 2020, accessed: 2026-01-14.

